# A Pattern Recognition Diagnostic Model to Restore and Emulate Knee Mobility

**DOI:** 10.1101/2021.12.23.21267314

**Authors:** David S. Hollinger, Muhammad Gulfam, Sadaira Packer, Bayu Sukmanto

## Abstract

Electromyography (EMG) is an electrical voltage potential linked to muscle contraction, resulting in human joint motion, such as knee flexion. Knee injuries, such as knee osteoarthritis (KOA), disrupt functional mobility of the knee joint and subsequently atrophy the muscles controlling knee movement during activities of daily living (ADL). Consequently, weakened muscles exhibiting deteriorated EMG signal fidelity are hypothesized to have discernible signal patterns from a healthy individual’s EMG signals. Pattern recognition algorithms are useful for mapping a set of complex inputs (EMG signals and knee angles) to classify knee health status (injured vs. healthy). A secondary outcome is to predict future knee angles from previous input signals to inform a robotic knee exoskeleton to apply real-time torque assistance to a patient during ADL. A Decision Tree Classifier, Random Forest, Naive Bayes, and a Feed Forward Neural Network (Fully Connected) were used for binary classification (healthy vs. injured). Partial Least Squares Regression, Decision Tree Regressor, and XGBoost were used to predict future joint angles for the regression task (knee angle prediction). Overall, the Random Forest Classifier had the best overall classification performance. XGBoost and Decision Tree Regression performed the best among regression algorithms for predicting real-time angles during walking while Partial Least Squares Regression performed the best during the standing tasks. In summary, our Machine Learning methods are useful for assisting clinicians and patients during physical rehabilitation by providing quantitative insight into the patient’s neuromuscular control of the knee.

## 1. Introduction

The estimated prevalence of limb loss in the US in 2006 was 1.6 million and is projected to grow to 3.6 million by the year 2050 [1]. Also, recurring knee pain affects approximately 25% of adults which limits function and mobility during activities of daily living (ADL) [2]. Neuromuscular traumas requiring rehabilitation to restore abilities to perform activities of daily living (ADLs) include spinal cord injury (SCI, 11,000 persons per year in U.S.), traumatic brain injury (TBI, 1.5 million per year), amputation (185,000 per year), and stroke (800,000 per year) [3]. There is a growing research effort towards building robotic assistive devices widely available for individuals with neuromotor deficit. Clinically realizable myoelectric control systems have been shown to enhance user control of a robotic limb. This control method uses the electrical muscle signal, known as electromyography (EMG), as input to control the robotic device. With the use of pattern recognition of the muscle signal, myoelectric-controlled powered exoskeletons can mimic a biological limb and assist individuals who struggle to perform ADL [4]. Therefore, the ability to predict healthy knee motion during common tasks, such as walking is essential for clinicians to monitor rehabilitation status among individuals with knee mobility deficits and for users to co-adapt with a robotic exoskeleton.

Clinical experts can potentially evaluate the analytic result of the proposed system to monitor patient outcomes in a rehabilitation paradigm. EMG signals are often too noisy and complex to create a generic musculoskeletal model to determine joint angles. Neural networks with non-linear activation functions have shown promise in mapping EMG signal inputs to joint angles. For example, Chan et al. showed success in identifying the complex mapping between full-wave rectified EMG signals and upper-limb trajectory using a Dynamic Recurrent Neural Network [5]. However, lower limb injuries outnumber upper limb injuries [1]. As a result, we decided to create a pattern recognition diagnostic to categorize the health status (injured vs. healthy) of an individual given their EMG activity and knee angle profile. The purpose of this study, therefore is twofold:

1. Classify an individual as injured or healthy based on EMG and knee angles
2. Predict knee angles of an individual at future time windows

## 2. Methods

### 2.1 Algorithms

Classification and regression algorithms were explored for predicting knee status (injured vs. healthy) and future knee angles. Binary classification evaluation was measured using classification accuracy (Eq. 2). Regression performance was measured using normalized root means squared error (NRMSE) (Eq. 1-3).

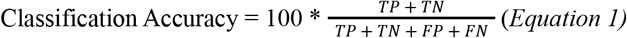

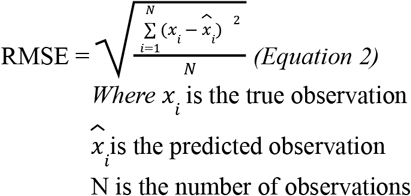

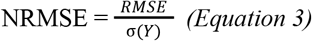

The supervised learning classifiers used in this study include a combination of simple and complex models. For example, a *Decision Tree* is a simple and versatile model with high interpretability. A Decision Tree starts at a root node and follows (at least) a binary decision to reduce the gini impurity score (Eq. 4).

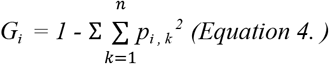

*where:*

*p*_*i,k*_ is the ratio of class k instances among the training instances in the i^th^ node.

Additionally, we used a *Random Forest Classifier* which is an ensemble learning algorithm composed of multiple decision trees, called forests. Random Forest is a powerful Machine Learning (ML) algorithm due to its robust ability to aggregate the predictions of many decision trees. See Appendix A for a list of the default parameters used as a baseline for Random Forest as well as the tuned hyperparameters used.

A *Naive Bayes Classifier* is a probability based method and is considered as a descent classifier even though as an estimator it is not considered very optimal. The core concept behind the naive bayes is conditional probability.

A Multilayer Perceptron or *Neural Network* method was also tested for the classification problem. We chose this model due to its established reputation in the deep learning community. A fully connected multi-layered perceptron was used for the experiment. One hidden layer was included in the architecture which includes dense layers with 100 hidden units. The neural network architecture is shown in Figure 1.

**Figure 1:**
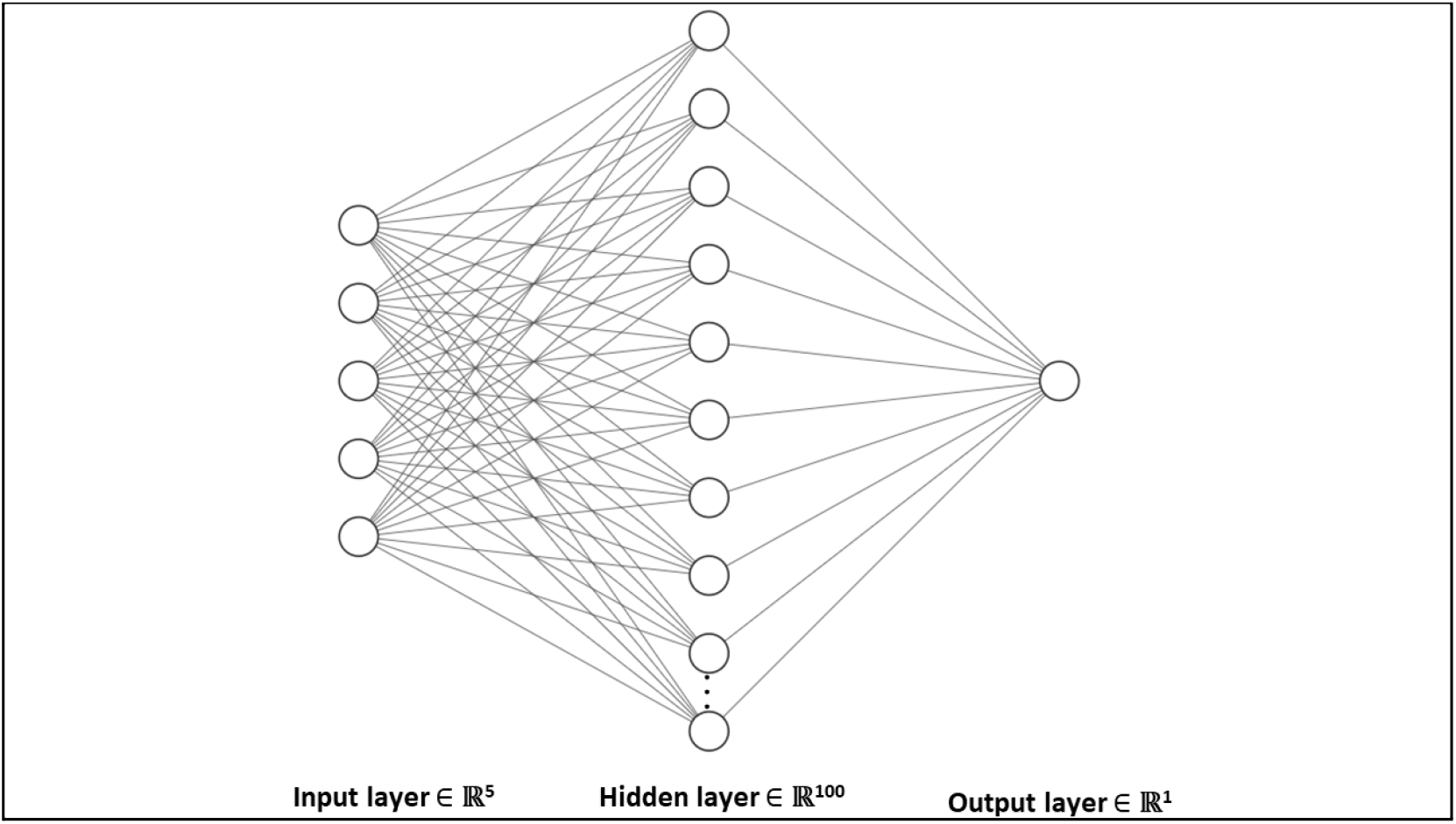
Neural network architecture with 5 input neurons, 100 neurons for a single hidden layer, and one neuron for the output layer

We trained and tested each model for walking, sitting, and standing. The results for each of the three exercises are reported in the results section. The Appendix A4 shows the training and validation accuracy for different training sizes.

For regression predictions, we included a diverse set of algorithms, such as Partial Least Squares Regression, Decision Tree Regression, and Extreme Gradient Boosting (XGBoost).

We decided to use a linear regression model for our knee angle prediction because it is a continuous dependent variable. We chose *Partial Least Squares Regression (PLSR)*. PLSR reduces the number of variables down to a smaller set of components and then performs least squares regression on the components. We also used a *Decision Tree Regressor* for the regression experiment because decision trees are capable of performing both classification and regression problems.

*XGBoost* is another powerful ensemble learning algorithm that is typically seen in Kaggle competitions due to its ability to produce high accuracy. XGBoost refers to the engineering goal to push the limit of computational resources for boosted tree algorithms by incorporating fast execution speed and high performance. The mantra of many Kaggle competition winners is, “When in doubt, use xgboost.”

For regression predictions, a rolling start cross validation (CV) method was used because temporal components of time series data require test set predictions to come after training data in a sequence (Fig. 2). Additionally, 80% of each CV split was used for training while the remaining 20% was used for testing. We chose to split each trial into 15 CV folds because it allowed the prediction window to be within 200 ms for testing.

**Figure 2:**
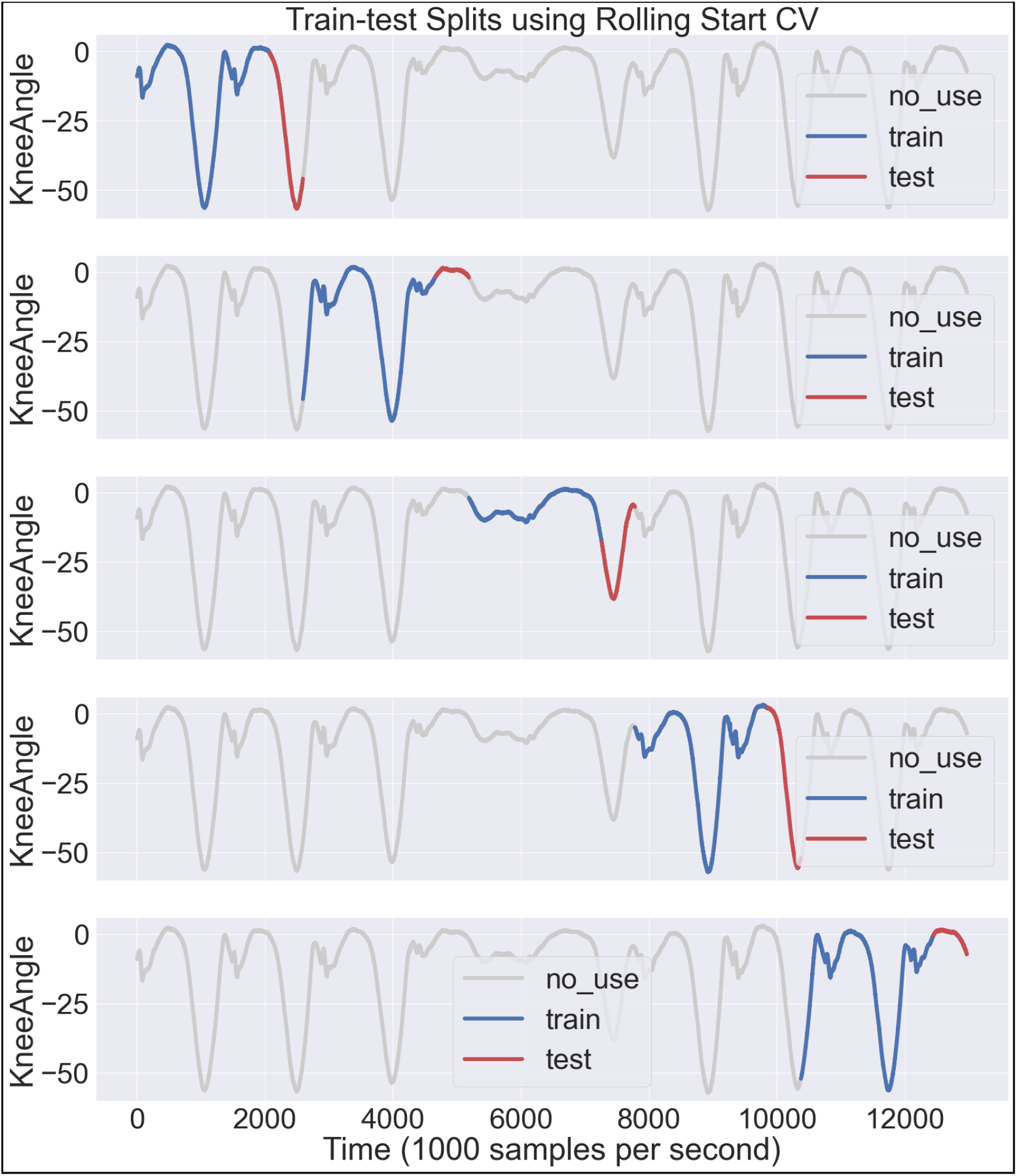
Example of rolling starting cross validation using 80% for training and 20% testing for each time window of a single subject.

### 2.2 Statistical Analysis

An Analysis of Variance (ANOVA) with repeated measures was implemented to observe the main effect between algorithms. Post hoc paired t-tests were performed when the main effect was statistically significant (p<0.05). Bonferroni corrections were also used during the post hoc test. All statistical analyses were performed in Microsoft Excel using the Analysis ToolPak..

## 3. Dataset

For this study, we accessed an open-source dataset [7] from the UCI ML Repository which included a multivariate time series data of five features (four EMG signal channels and one knee angle) (Fig. 3). The data was collected using electromyography (EMG) electrodes to obtain muscle activation signals typically occurring when a person flexes and extends the knee. The three movements include leg extension from a sitting position, knee flexion from a standing position, and normal walking (Fig. 4). The EMG electrodes were attached to four muscles: biceps femoris, vastus medialis, rectus femoris and semitendinosus while a goniometer sensor measured knee angles (Fig. 5).

**Figure 3:**
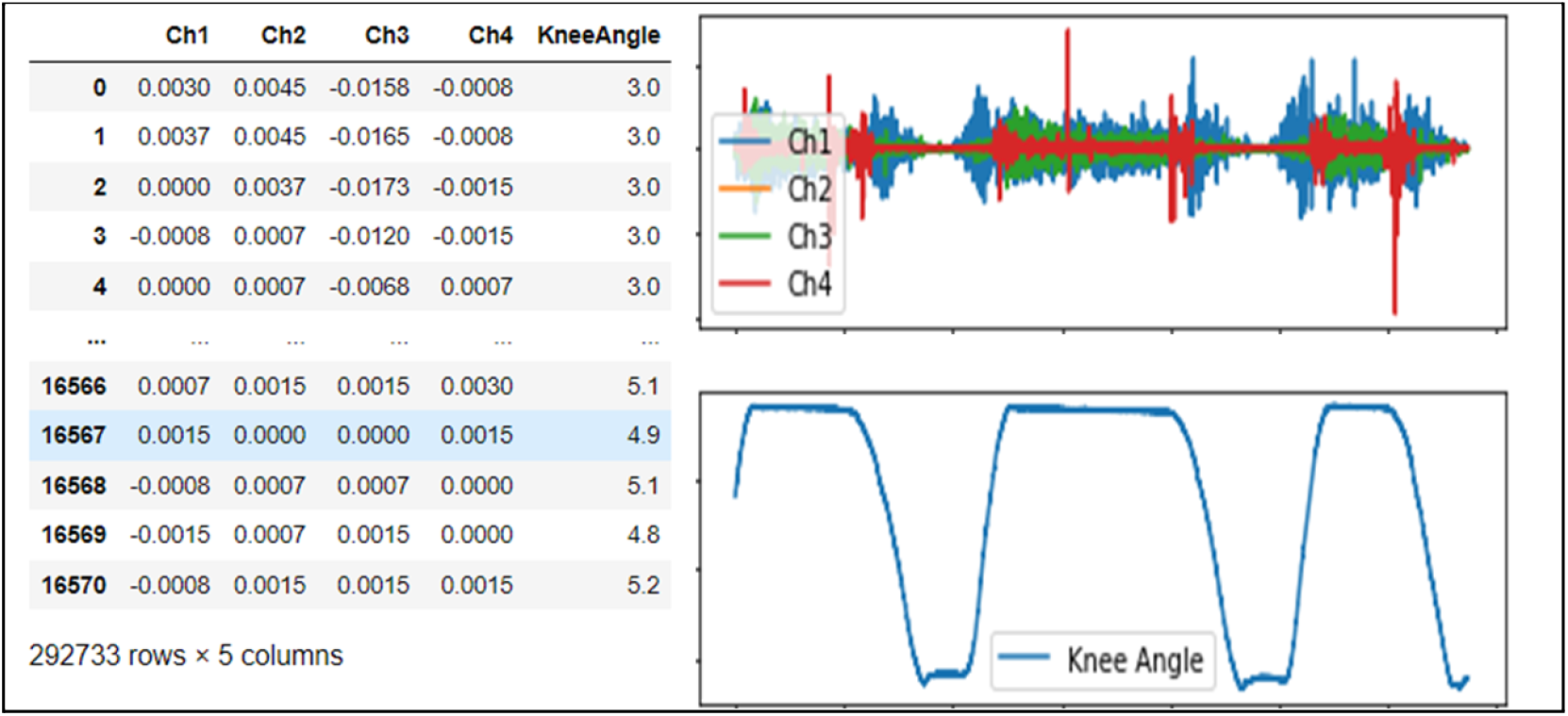
Multivariate time series from open-source Machine Learning Repository.

**Figure 4:**
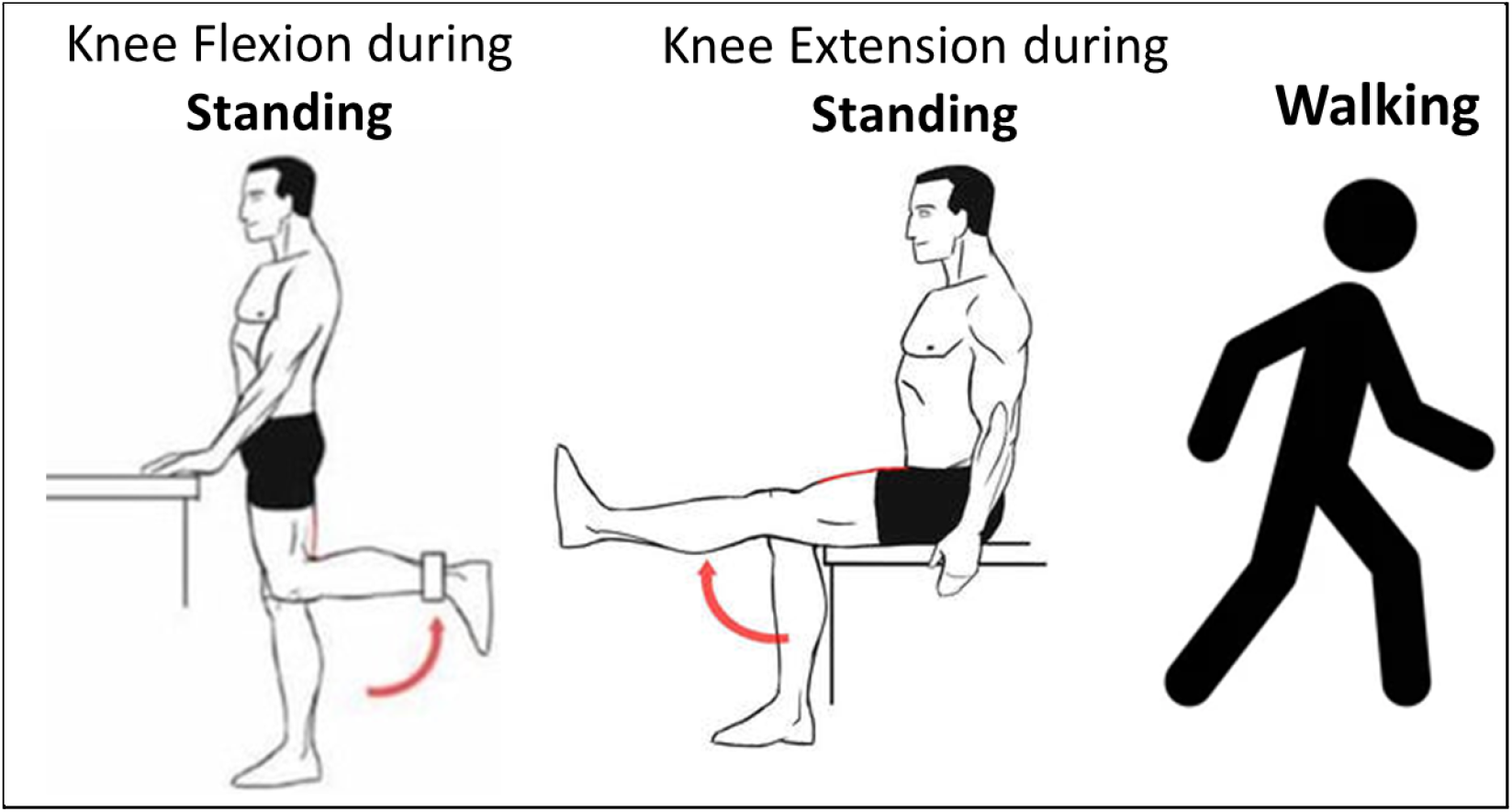
Actions corresponding to knee motion performed by each subject during data collection

**Figure 5:**
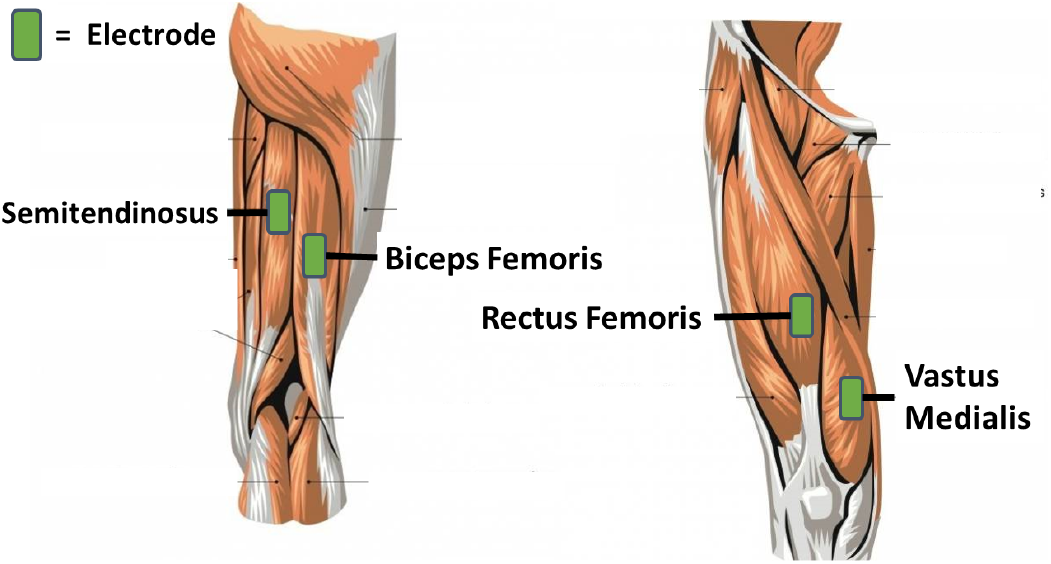
Electrode for measuring EMG signal for the knee flexor and extensor muscles.

The first experiment compared the classification performance of different algorithms predicting knee injury status (healthy vs. injured). For the second experiment we compared regression models for predicting the motion of the knee angle. The binary classification is important because of the utility of diagnosing a subject specifically on the knee because of the subject’s deteriorating health condition in relation to the knee angle during an examination.

The data is collected from 22 participants from which 11 were normal participants and the other 11 were diagnosed with knee abnormalities. The data was collected for three different types of activities which include walking, leg extension from a sitting position, and flexion of the leg up.

The foreseen measurement, as the result of the evaluation, is based on classification accuracy of healthy vs. impaired knees. For the regression prediction, the model was evaluated using root mean squared error (NRMSE).

### 3.2 Data Preparation

The features for this multivariate time series included four columns of EMG channel signals and one column of knee angles (Fig. 6). Classification tasks incorporated five columns (4 EMG + 1 Knee Angle) as input features whereas regression used four EMG channel columns as feature inputs to predict knee angles. Additionally, 10-fold CV was used to reduce the likelihood of overfitting a model during classification.

**Figure 6:**
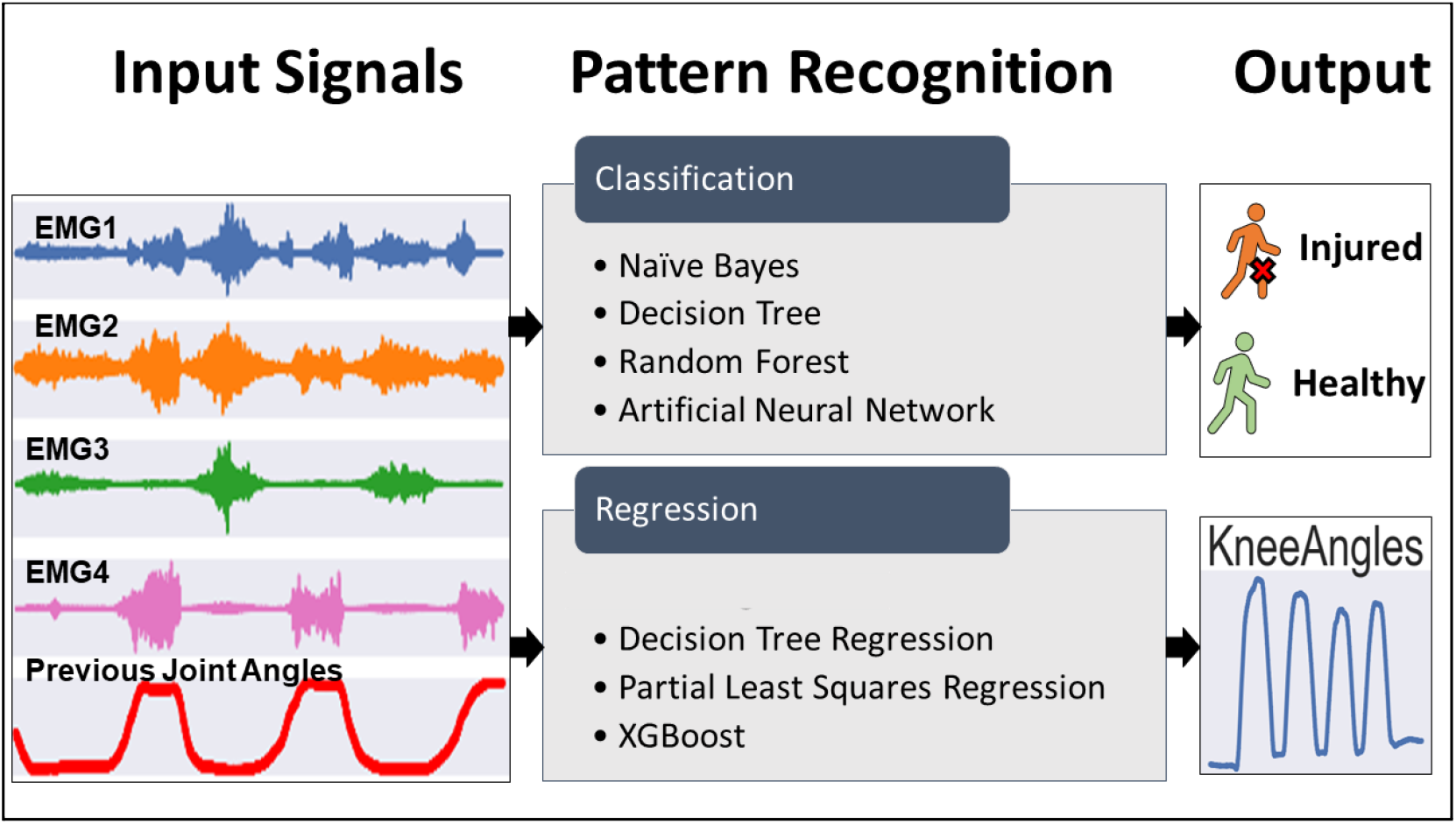
Flowchart of the input signals, algorithms, and outputs used for predicting the health status (classification) and joint angle (regression) of a patient.

## 4. Experiments

The hyperparameters chosen for the classifiers and regression algorithms mentioned in the methods section are derived from the default values from Scikit-learn’s machine learning library. The metrics chosen were classification accuracy for the classification task and normalized root mean squared error (NRMSE) for the regression task (Eq. 1-3)

For the Naive Bayes Classifier, default parameters were used to train the Naive Bayes Classifier which include priors (default=“none”) and var_smoothing (default=1e-9). The results for Naive Bayes Classifier are reported in the results section.

Besides mentioned models in the above paragraphs, the feedforward neural network included a single hidden layer with the size of 100, relu as the activation, adam as the solver chosen, and alpha parameter of 0.0001. As the decision tree is also used in this work, most of the configuration is in default setting from Scikit-learn’s ML library in Python. The Decision tree regressor uses 0.0 ccp alpha, mse as the criterion, 0.0 minimum impurity decrease, 1 minimum samples leaf, 2 minimum samples split, 0.0 minimum weight fraction leaf, best as the splitter, and none setting for maximum depth, maximum features, maximum leaf nodes, minimum impurity split and random state.

Further, for the partial least squares regression model (PLSR), the model uses two components, with scaling, across 500 iterations, with copy enabled, and 1e-06 as the tol. As this work uses XGBoost too, the model is configured to use squared error regression, 0.5 base score, gbtree as the booster, 0 gamma, -1 gpu ID, learning rate of 0.300000012, 0 maximum delta step, 6 maximum depth, 8 number of jobs, 1 number of parallel tree, 0 random state, 0 regression alpha, and exact tree method. While for the other parameters like columns sample by level, column sample by node, columns sample by tree, minimum child weight, number of parallel tree, regression lambda, scale pos weight, subsample, and validate parameters are all set as 1.

All models used in this work learn from the same data set arrangement as previously explained in the data set and data preparation sections. The data contains information taken from muscles and knee flexion as explained on figure 5. Within the data, field numbering corresponds to the same numbering of the device sensor channels. Rectus femoris, biceps femoris, vastus internus semitendinosus and knee flexion sensed data are put into the field 0, 1, 2, 3, and 4 through channel 1, channel 2, channel 3, channel 4 and channel 5 respectively. Those data fields are found in every single file within the data set that represents a task of a subject (sitting, walking, or standing) with specific status (normal or abnormal).

The data then being partitioned into training set and test set as explained on figure 5. During a series of model learning, the bottom line of the model, that is data class balancing, was found to be compulsory for improving the learning accuracy. The result section will explain more about the data class imbalance issue firstly found and how it impacts the learning result. The population differences due to the imbalance issue could be shown on figure 7. As this issue appears, the experiment is then being rolled back to the data preprocessing as well as the dimensionality reduction. For further mitigation during the experiment (such as for overfitting risk), k-folds cross validation with the k of 10 is needed with the result sampled on figure 6. Every k uses a data split of 80% and 20% for training and testing respectively. An improved result of the cross validation approach could be shown on figure 10.

**Figure 7:**
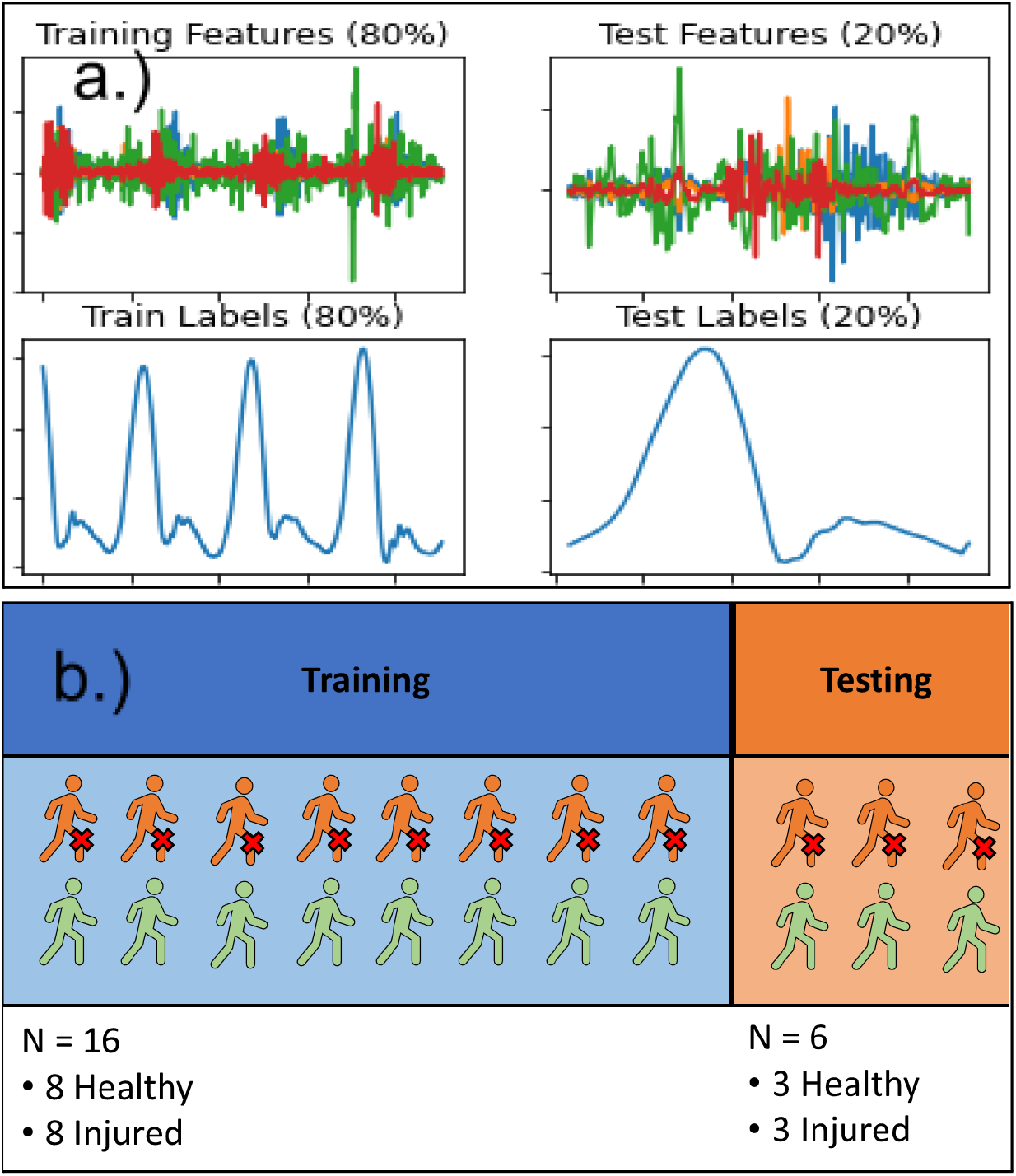
Training and testing sets for a.) regression. b.) classification

**Figure 7:**
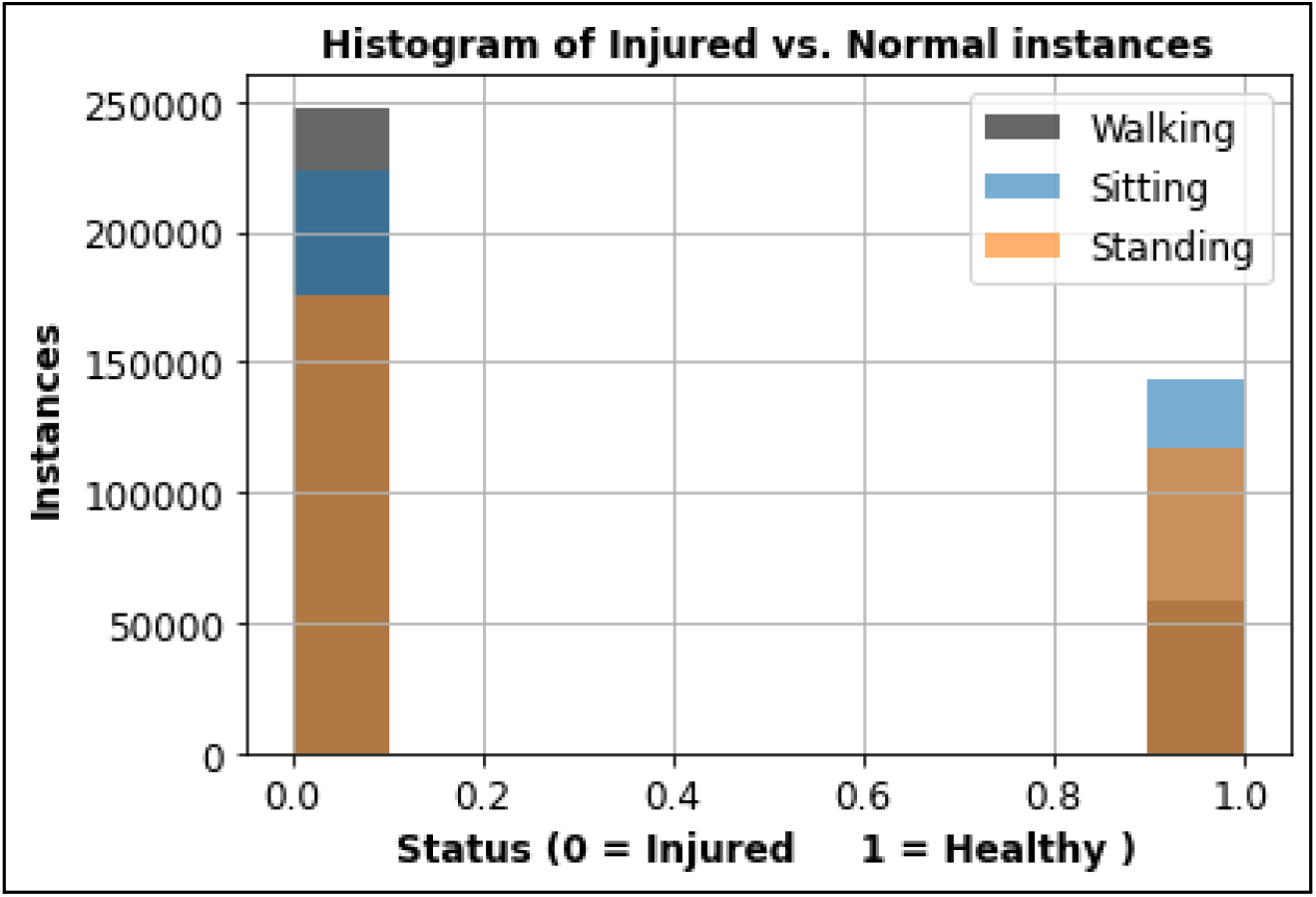
An example of class imbalance shown in the training set for each task.

Various graphs are used for the data visualization such as histogram, bar charts, boxplot, correlation heatmap, and time series curve. Not only for showing the learning result and comparing results of every model at every learning iteration, but it could be useful as well during the data preprocessing as shown on figure 8 that identifies the existence of possible outliers within the data set.

**Figure 8:**
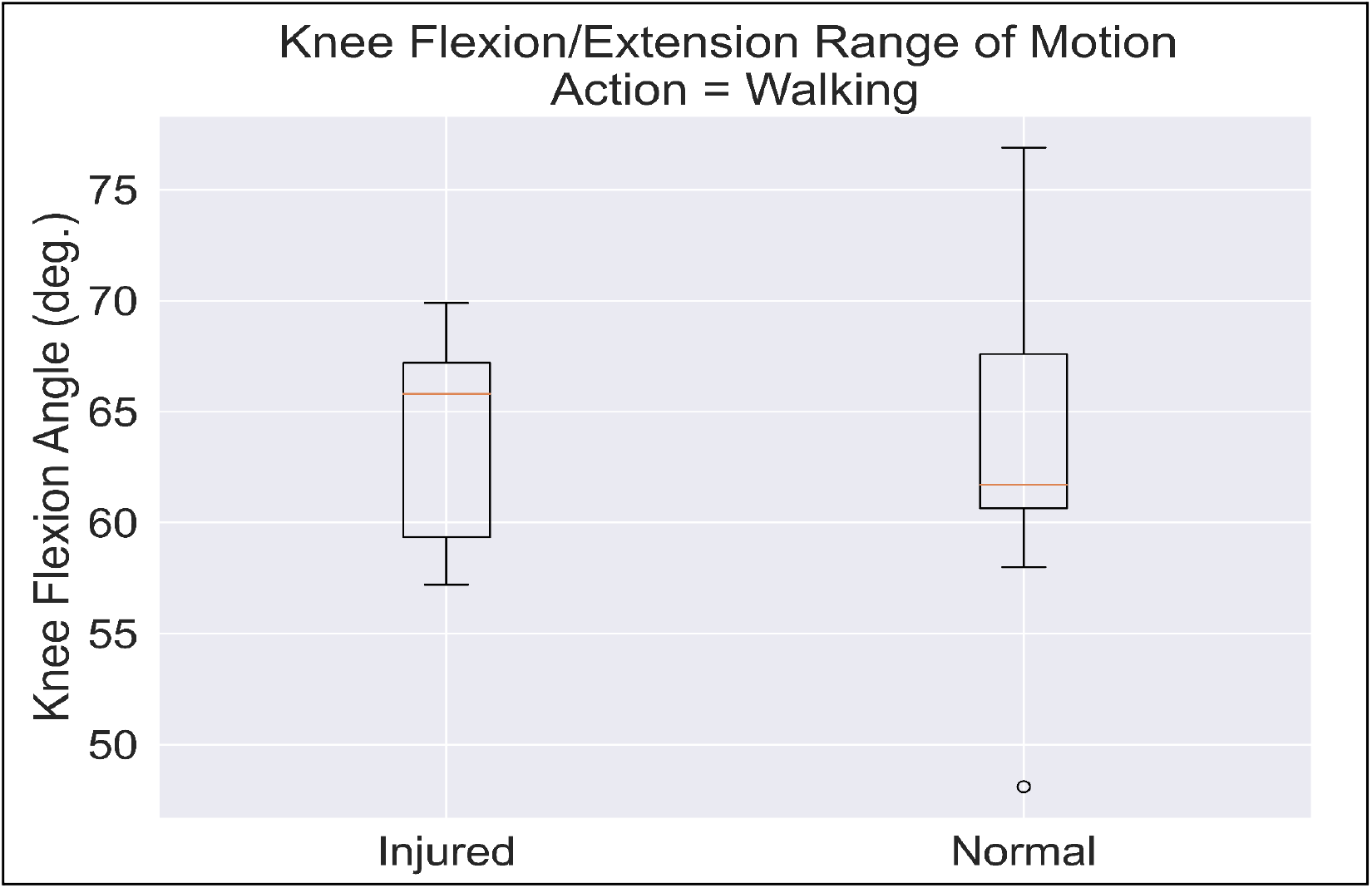
Knee range of motion for 11 injured and 11 healthy participants during walking

The models used for learning the data are explained on figure 6. For implementing classification and regression, we used a Python Jupyter notebook because there are a plethora of powerful ML models available in the scikit learn library. The method section already explains how each model works and what metric is used to measure the models. To compare the classification accuracy as well as regression metrics (NRMSE), the comparison table, like tables 1-3 respectively, shows accuracy parameters for each classification model at each activity (sitting, standing, and walking).

**Table 1:** Classification results for walking, sitting, and standing tasks. The best performance for each task is in **bold font**.

| Method | Accuracy (%)<br>Walk | Accuracy (%)<br>Sit | Accuracy (%)<br>Stand |
| --- | --- | --- | --- |
| Naive Bayes | 75.54 | 63.23 | 66.10 |
| Decision Tree | 85.81 | 75.95 | 69.37 |
| Random Forest | 89.23 | 80.11 | 75.64 |
| Neural Network | 89.36 | 77.65 | 71.05 |

**Table 2:**
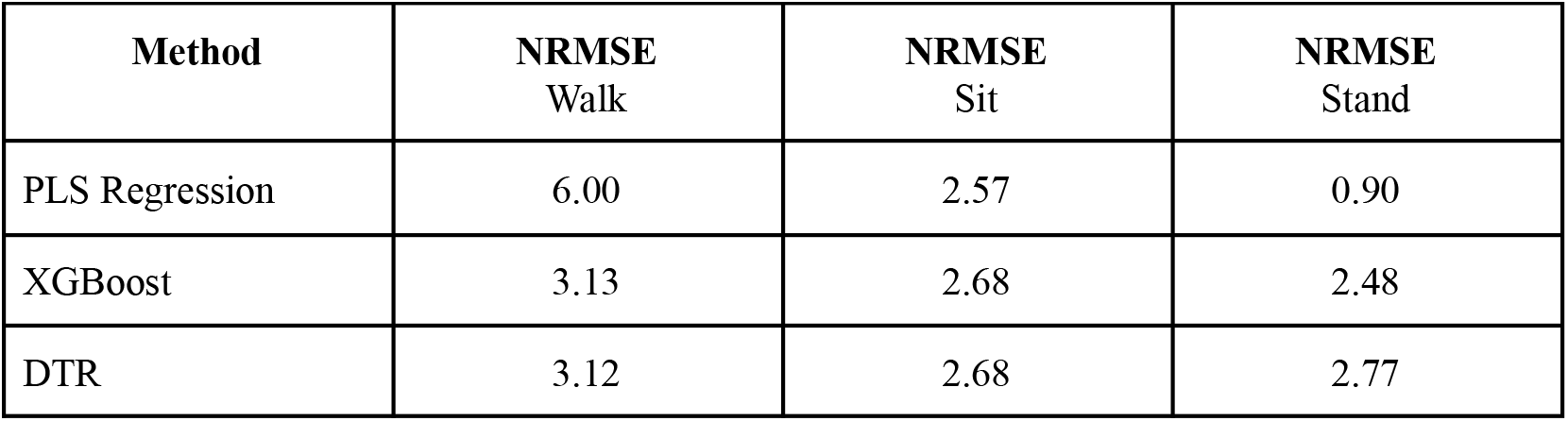
Prediction performance for regression algorithms during walking, sitting, and standing

**Table 3:** Mean NRMSE for regression models during walking, sitting, and standing tasks. **Bold** result indicates statistically significant results for ANOVA and for paired t-test during post hoc test with a Bonferroni correction.n.s. indicates a non-significant result.

| **<u>ANOVA Result (p-value)</u>** |  |  | **<u>Paired t-test (p-value)</u>** |  |  |
| --- | --- | --- | --- | --- | --- |
| Walking | Sitting | Standing | PLSR vs. XGBoost | PLSR vs. DTR | XGBoost vs. DTR |
| **1.2E-10** | 0.93 | **0.000011** | **1.46E-08** | **1.37E-08** | 0.98 |
|  |  |  | n.s. | n.s. | n.s. |
|  |  |  | **0.00018** | **0.000020** | 0.43 |

By conducting all iterations of this work, with improvement at each iteration, we would finally expect to show which learning model that best fits this particular problem. By choosing two fundamentally different models, simple and complex, it would increase the chance to decide the best learning model.

## 5. Results

A data exploration pipeline was performed prior to testing model performance. The data was not previously preprocessed so we tested the data for class imbalance (Fig. 7) and dimensionality reduction (Fig. 9). Additionally, the knee range of motion was computed for injured and healthy patients to observe whether the values correspond to typical values seen in literature (Fig. 8). As shown in the boxplots in Figure 8, the healthy group surprisingly had a higher standard deviation of range of motion which is different from previous studies. However, the values were not drastically different (>10 deg.) to confidently throw out the data [6].

**Figure 9:**
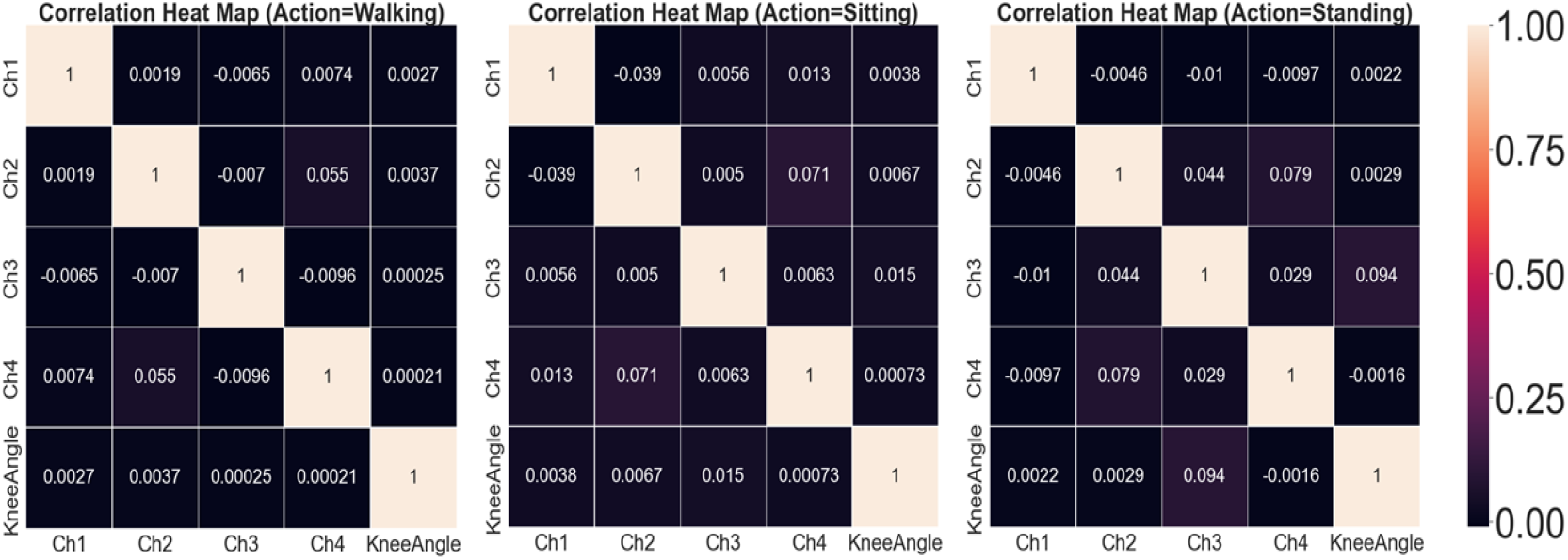
Correlation heatmaps for walking, sitting, and standing. The features do not have multicollinearity so feature reduction was not needed.

Knee range of motion is a crucial outcome metric following injury and is typically assessed following knee arthroplasty [6]. Therefore, we observed the knee range of motion for each subject to make sure the training and test sets contained a balanced distribution of subjects with comparable knee mobility (see Figure A6 in Appendix A).

On a comparatively balanced dataset we tested our neural network method and obtained the accuracy of 89.36%, 77.65%, 71.05% for walking, sitting, and standing exercises, respectively (Table 1, Figure 10). The results for all classification and regression algorithms are shown in Tables 1-2. The prediction accuracy for the different exercises data shows that walking data is a complex movement that allows for greater discriminability power.

**Figure 10:**
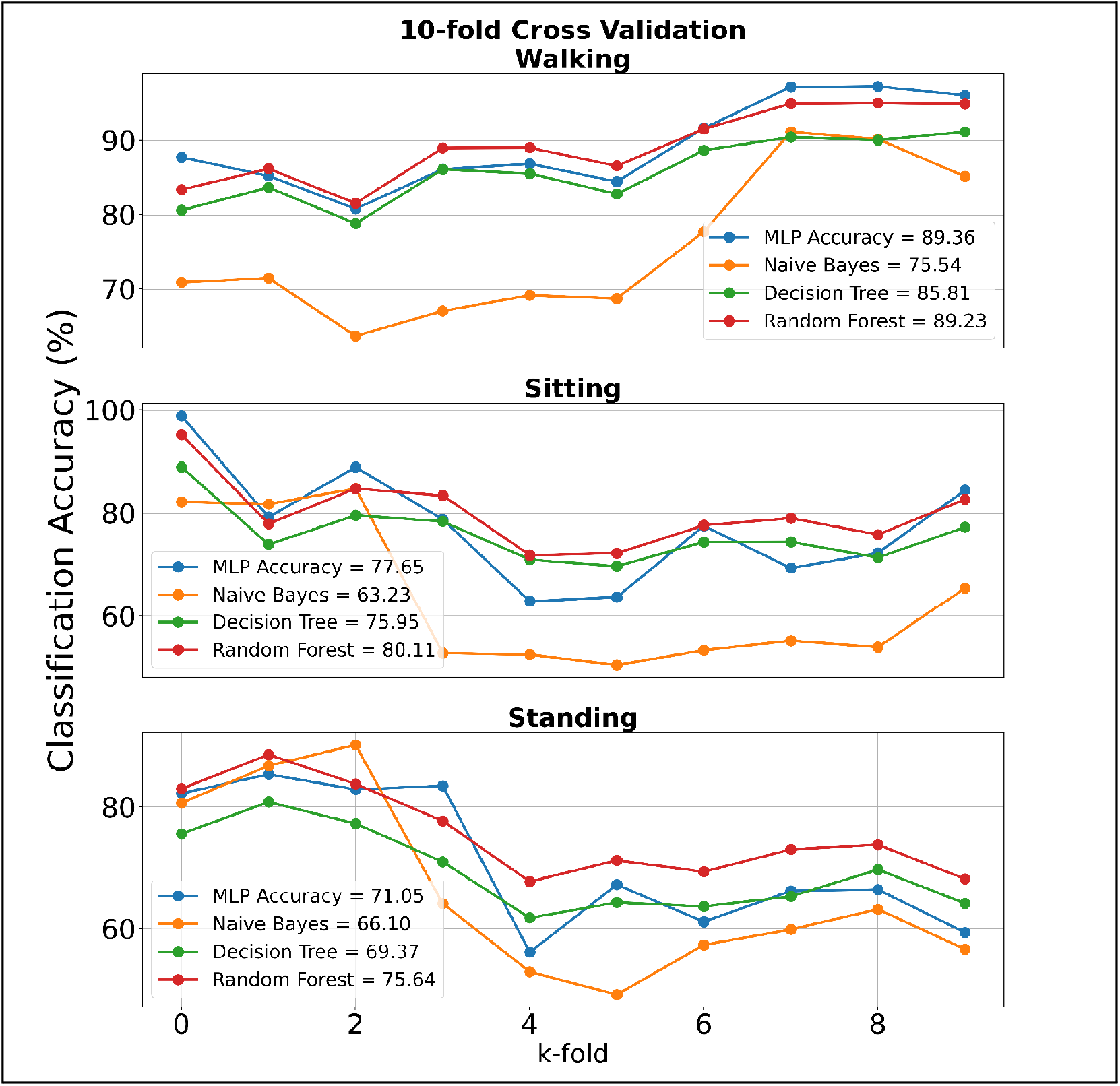
Classification accuracy for supervised learning algorithms during walking, sitting, and standing actions.

**Figure 11:**
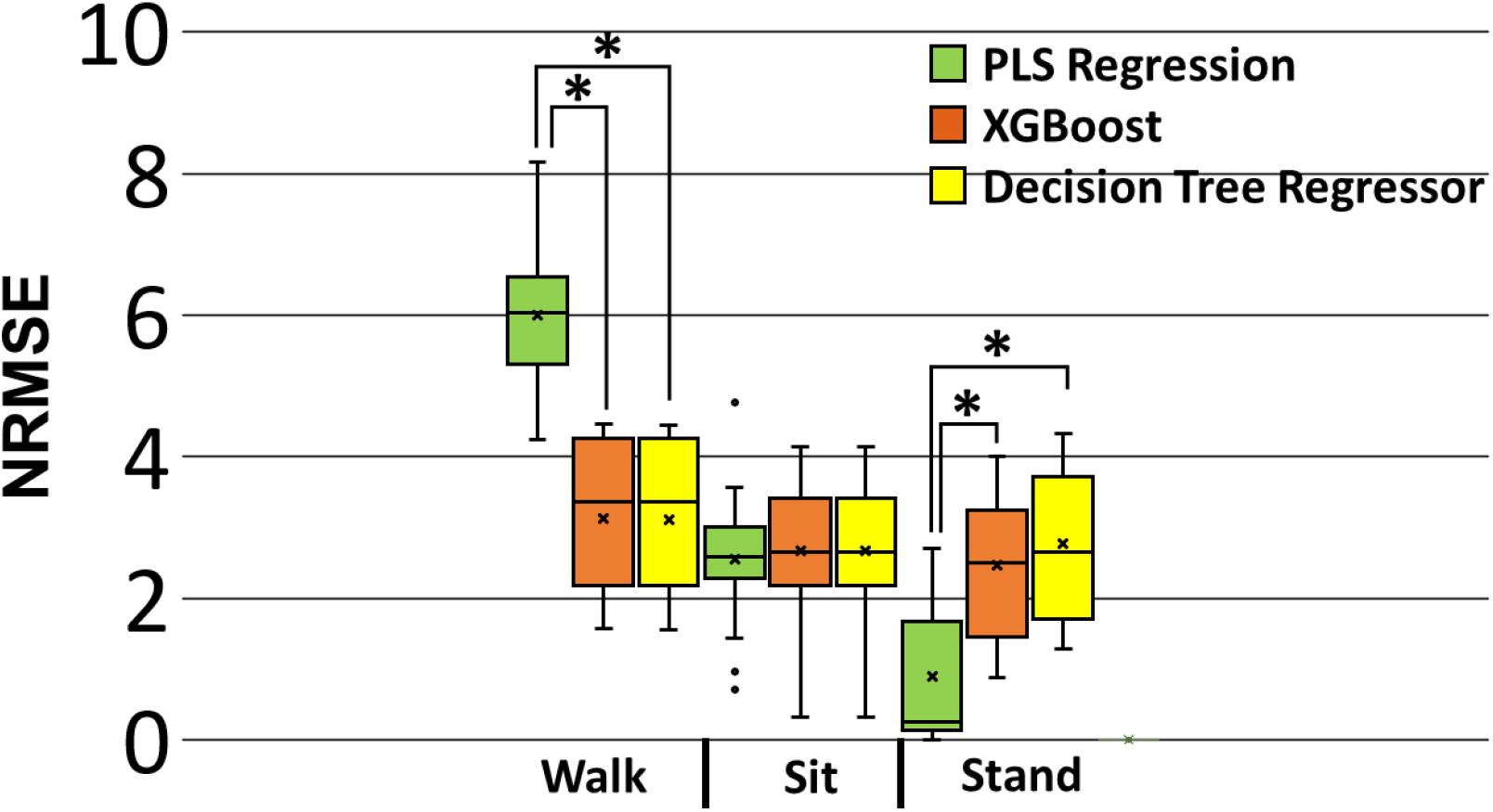
Comparison of regression algorithms for predicting knee flexion angle during walking, sitting, and standing tasks.*indicates statistically significant comparison (p<0.05).

## 6. Discussion

There are several key takeaways from using pattern recognition algorithms of EMG signals to determine the health status of the knee. These key findings are:

### 6.1 Class imbalance in training and test sets decreased model generalizability

Prior to performing the data exploration pipeline for the balanced dataset, accuracy was considerably worse. Classification accuracy from the imbalanced datasets was around 50%, which is not better than random chance. Balancing the dataset improved performance slightly by a classification accuracy of 2-3%. Although, using CV methods showed to be more useful to select a balance of injured and healthy participants. We made sure the CV included time-series relevant predictions by setting the parameter Random Shuffling = False.

### 6.2 Complex models outperformed simpler models for binary classification

The accuracy for Random Forest (89.23%, 80.11%, and 75.64%) and Neural Network (89.36%. 77.65%, and 71.05%) outperformed Naive Bayes and Decision Tree Classifier. This result shows the powerful performance capability of the ensemble method from Random Forest is useful when observing complex time series data, such as EMG. Additionally, a Neural Network of only one hidden layer is nearly 90% accurate for determining the injury status of an individual using signals from walking data. Further development of a Recurrent Neural Network (RNN) and Long Short-Term Memory (LSTM) could improve prediction performance because the neurons in the RNN and LSTM loop in both directions, therefore enhancing the ability to harness the temporal aspects of the features. A feedforward Neural Network, on the other hand, only allows the neurons to travel in one direction.

### 7.3 Classification models performed best during the walking task

All classification models used performed best during the walking task. In essence, walking tasks allow for more easily discernible muscle signals that are distinguishable between injured and healthy populations. The sitting and standing task only used a single degree of freedom movement (knee flexion or extension) which is more difficult for algorithms to discriminate between healthy and injured groups. Consequently, clinicians should incorporate tasks for patients that involve multiple degrees of freedom, such as walking.

While improving the models used within this work, one of the most valuable findings was about class imbalance in the data that can lead to many problems like high accuracy but useless models. In the real world while examining subjects with injured joints, like the knee, observers tend to take a longer duration compared to healthy subjects. This might be due to ensuring the judgment of subject abnormality. Data preprocessing for balancing this data set is needed prior to retraining the model. We found that using a CV of the training and test sets resulted in improved accuracy and generalizability. Originally, the models had trouble ‘learning’ the relationship between the train/test sets since we hard coded the training & testing set (Fig. 7). In other words, the first 8 subjects were not a good training set to learn a relationship between feature inputs and output labels to generalize the last 3 subjects. CV using 10 folds ensures the models fit to a more generalized combination of training sets, therefore improving the generalization on the test set.

For predicting the pattern of the multivariate time series data, some regression models are used in this work. The result of the DTR model is comparable with the XGBoost for each of the three tasks. However, PLS Regression performed significantly better during the standing tasks and significantly worse during walking tasks. This may be due to the fact that XGBoost is better able to predict the complex walking task compared to PLS Regression.

There are a few limitations and future work worth mentioning. We realize that learning multivariate time series data will remain relevant as the development of the algorithms, improved generalized models, and broader domains in reality. The challenging approach of this work is due to the multivariate time series data to find the best way to analyze the data in a seamless pipeline. For example, different kinds of knee injury may lead to different patterns of multivariate time series data. We hope this work will motivate readers to further the journey of introducing pattern recognition models to the health of either a system or other biomechanical applications.

## 7. Conclusion

Pattern recognition algorithms can open the door to the possibility of a knee diagnostic for clinicians to monitor their patients rehabilitation outcomes. Further development using hyperparameter tuning can increase prediction accuracy and possibly reduce computational speed. This study is one of many ongoing projects using multivariate time series data collected from worn sensors to advance human performance.

## Data Availability

All data produced in the present study are available upon reasonable request to the authors.

http://archive.ics.uci.edu/ml/datasets/emg+dataset+in+lower+limb#

## Acknowledgments

A special thanks to Gerry Dozier for providing general feedback on the structure of the paper and to Bo Liu for algorithm recommendations.

## 5 APPENDIX A

Architecture for each model used:

**Figure A1:**
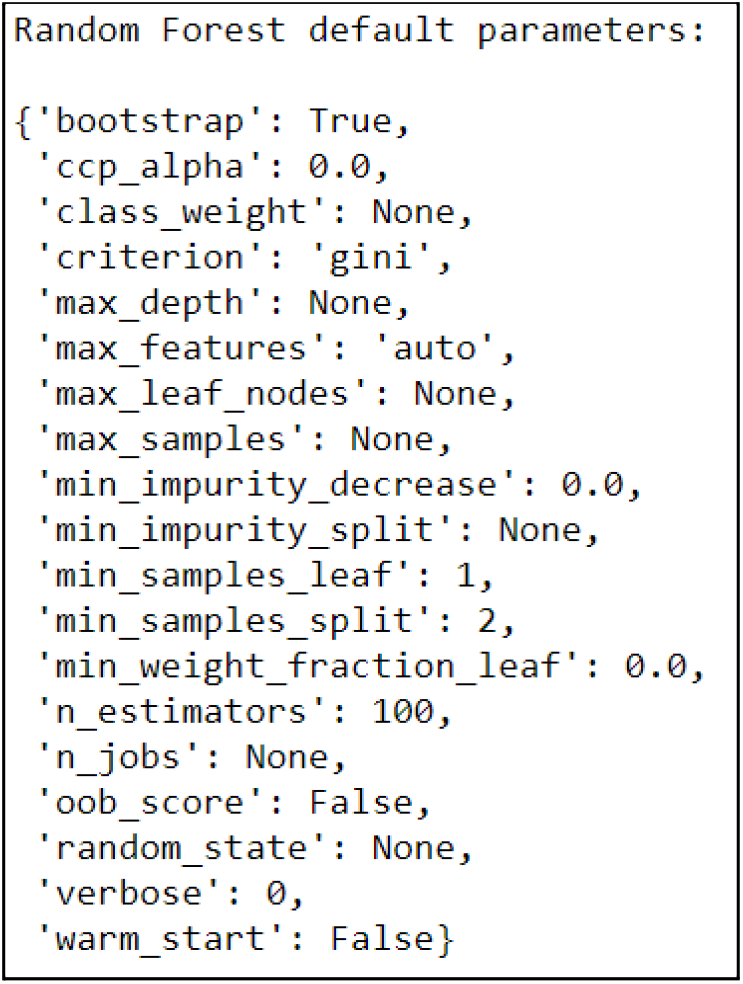
Baseline Random Forest parameters from the default values in sklearn library:

**Table A1:** Neural Network Architecture

|  |  |
| --- | --- |
| Hidden Layers | 1 |
| Hidden Units per layer | 100 |
| Solver | Adam |
| L2 penalty (regularization term) parameter | 0.0001 |
| learning_rate | 0.001 |
| max_iter | 200 |

**Figure A2:**
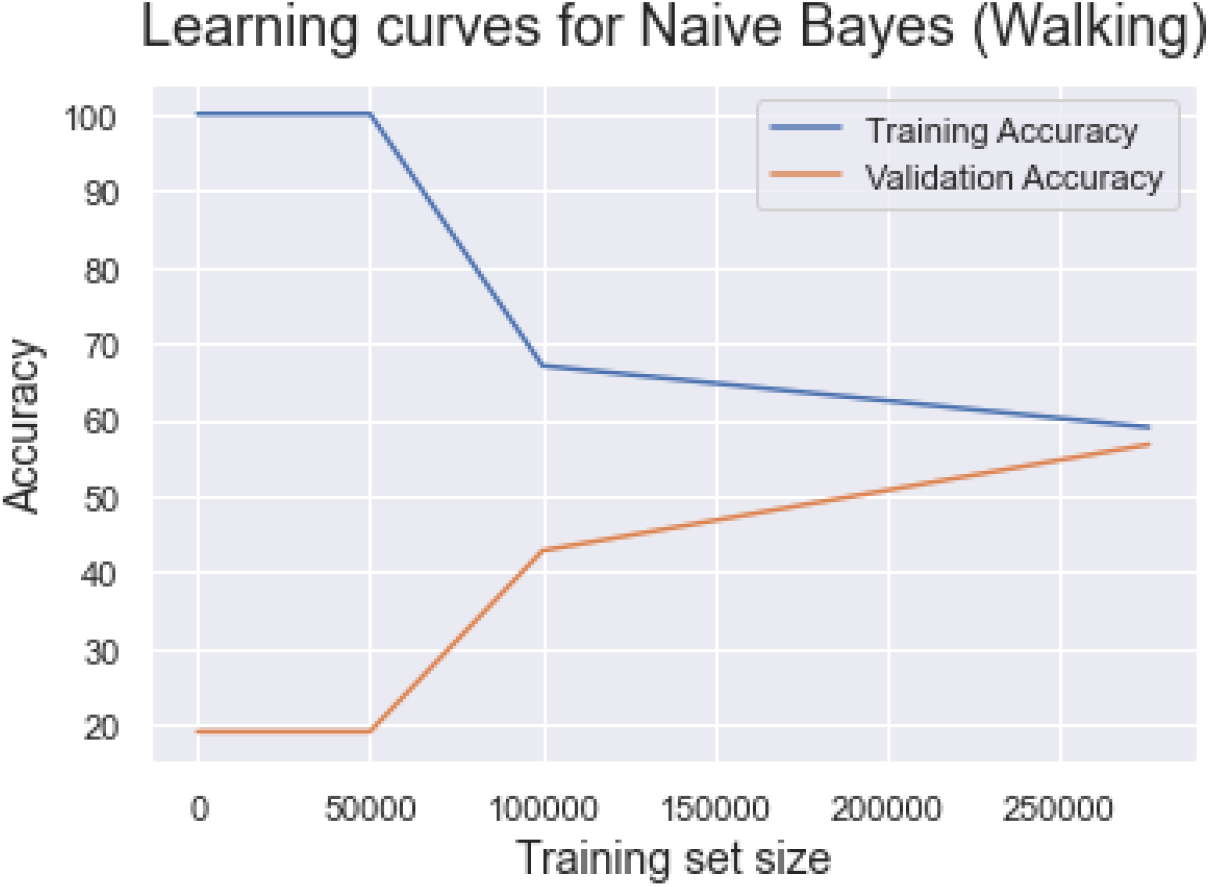
Training and validation accuracy for Naive Bayes. Train_sizes has been interpreted as absolute numbers of training samples and must be within (0, 275688]. We, therefore, expect higher validation accuracy as the training size maximum = 306320.

**Figure A3:**
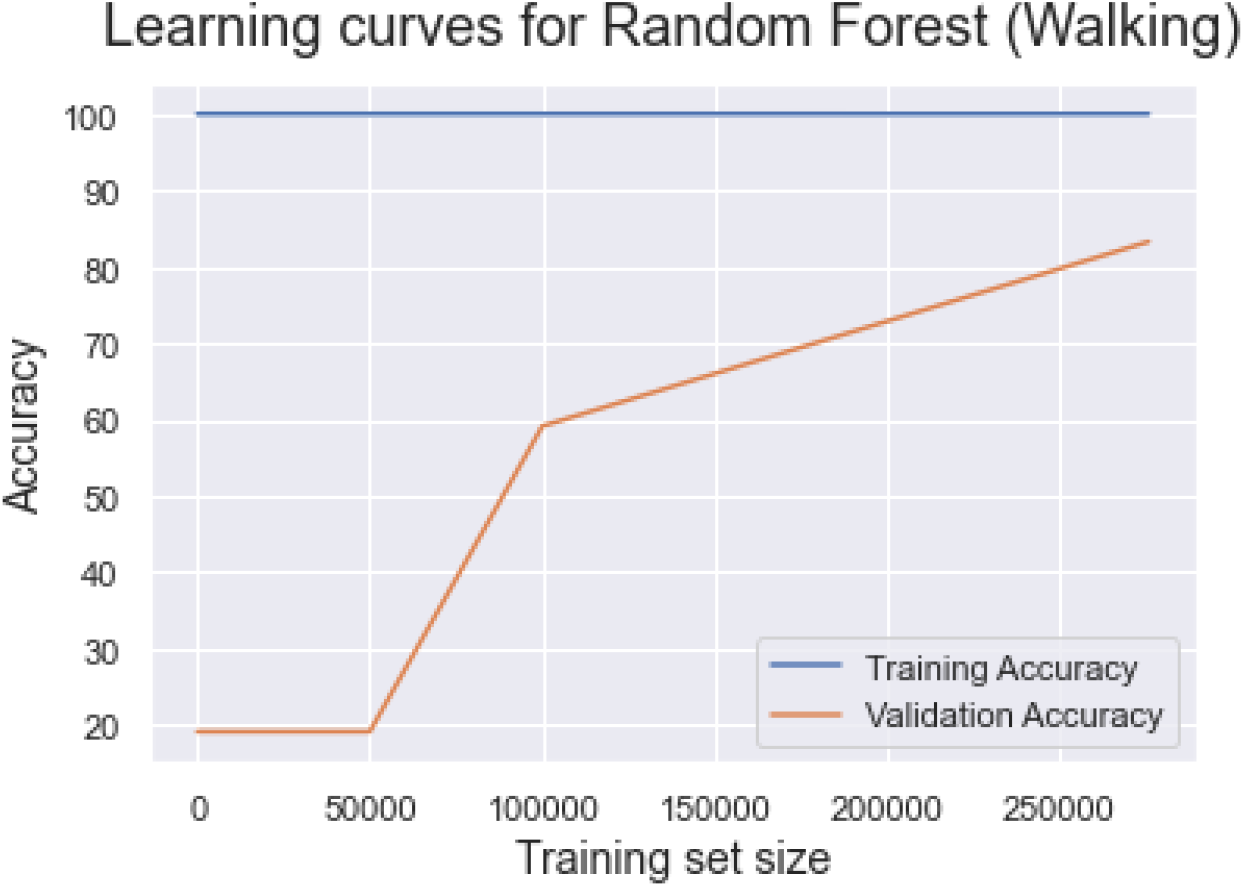
Training and validation accuracy for Naive Bayes.

**Figure A4:**
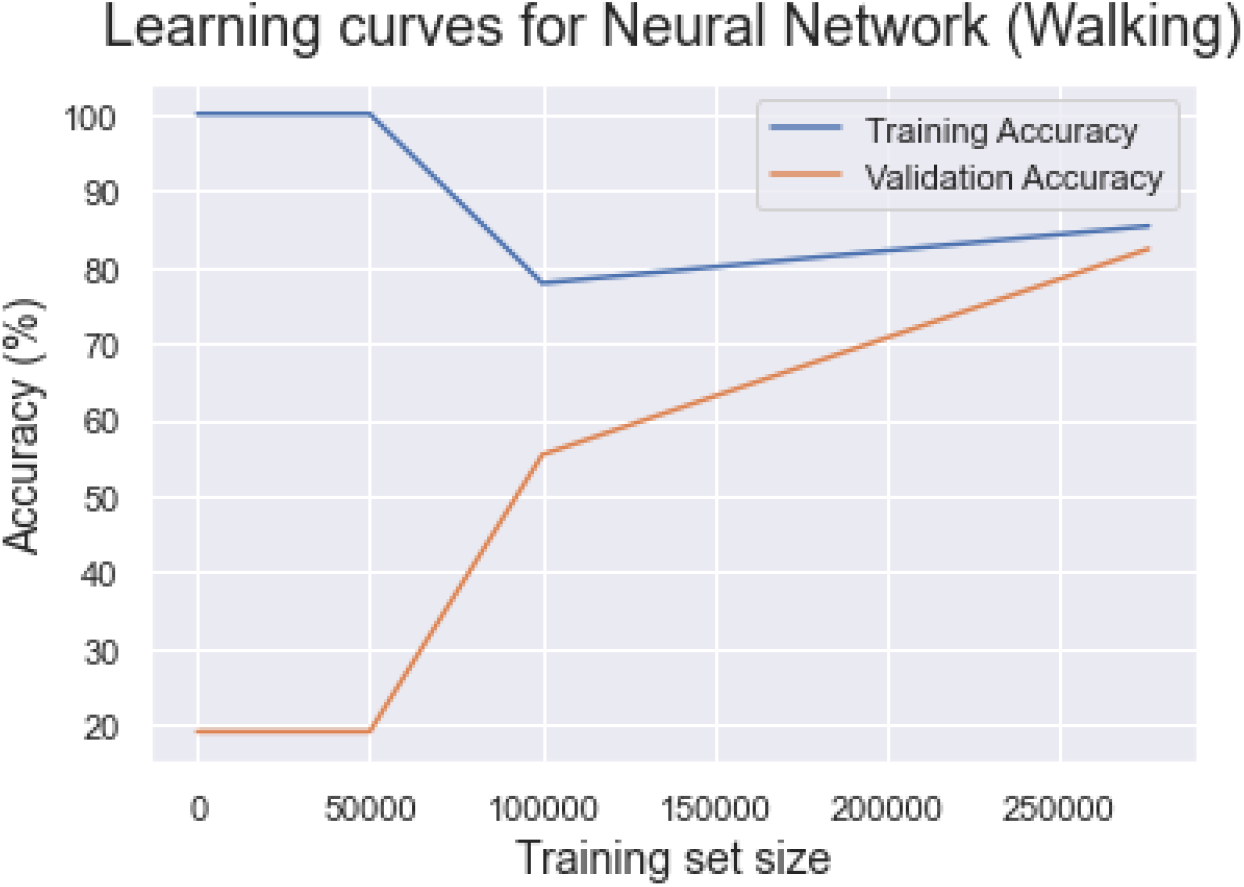
Training and validation accuracy for Neural Network.

**Figure A5:**
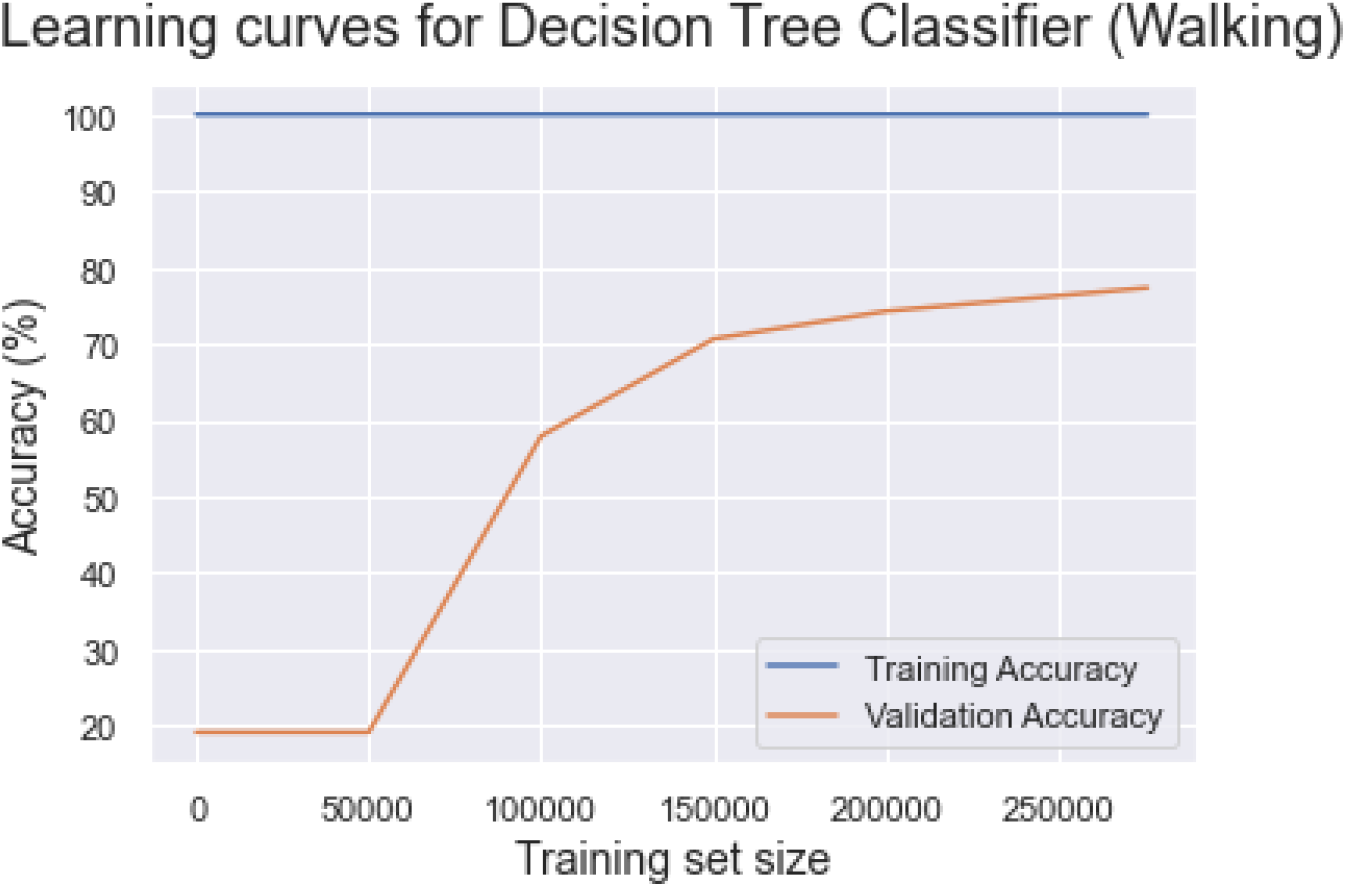
Training and validation accuracy for Decision Tree Classifier

**Figure A6:**
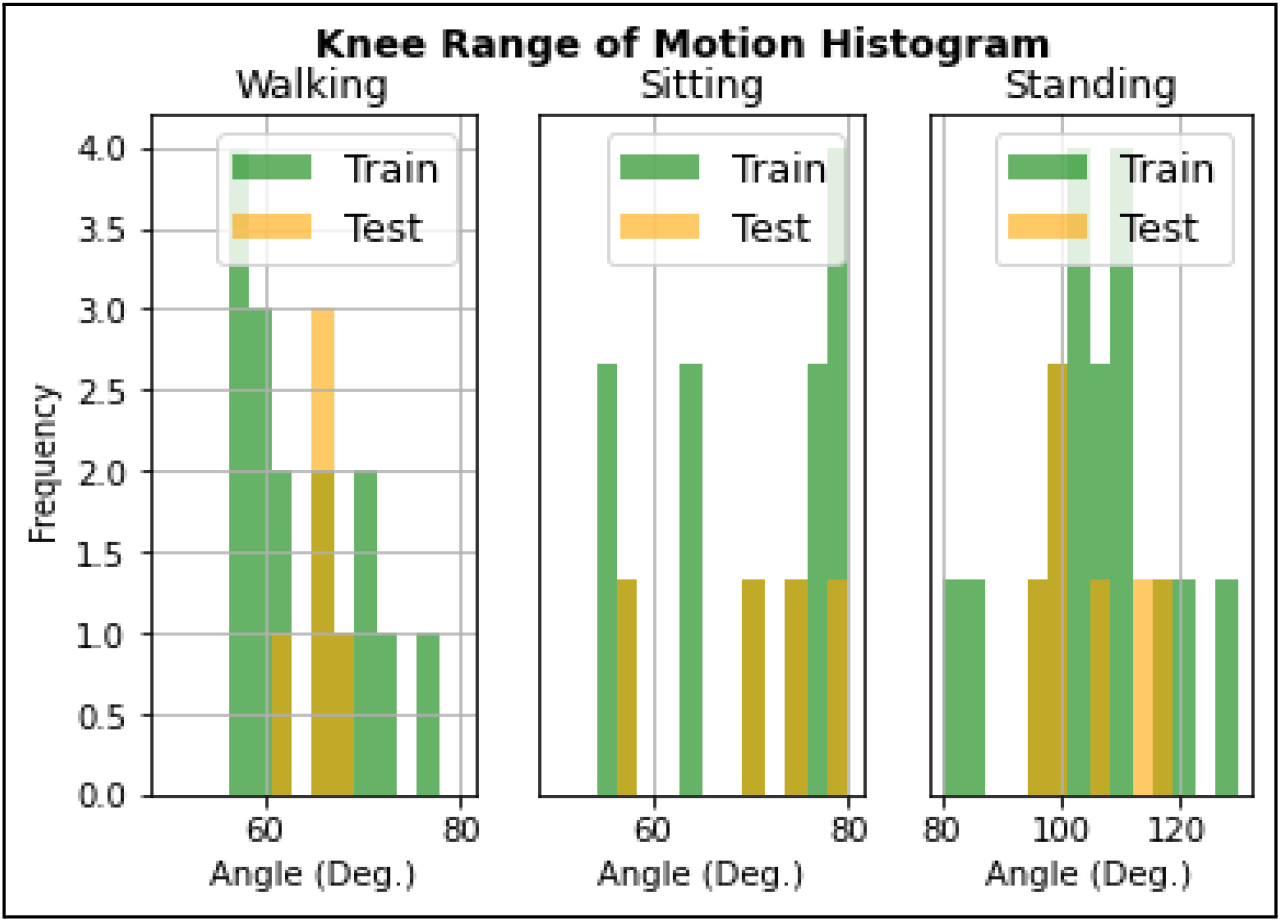
Knee Range of motion for each of the 22 subjects in the study in the training and test sets.

**Figure A7:**
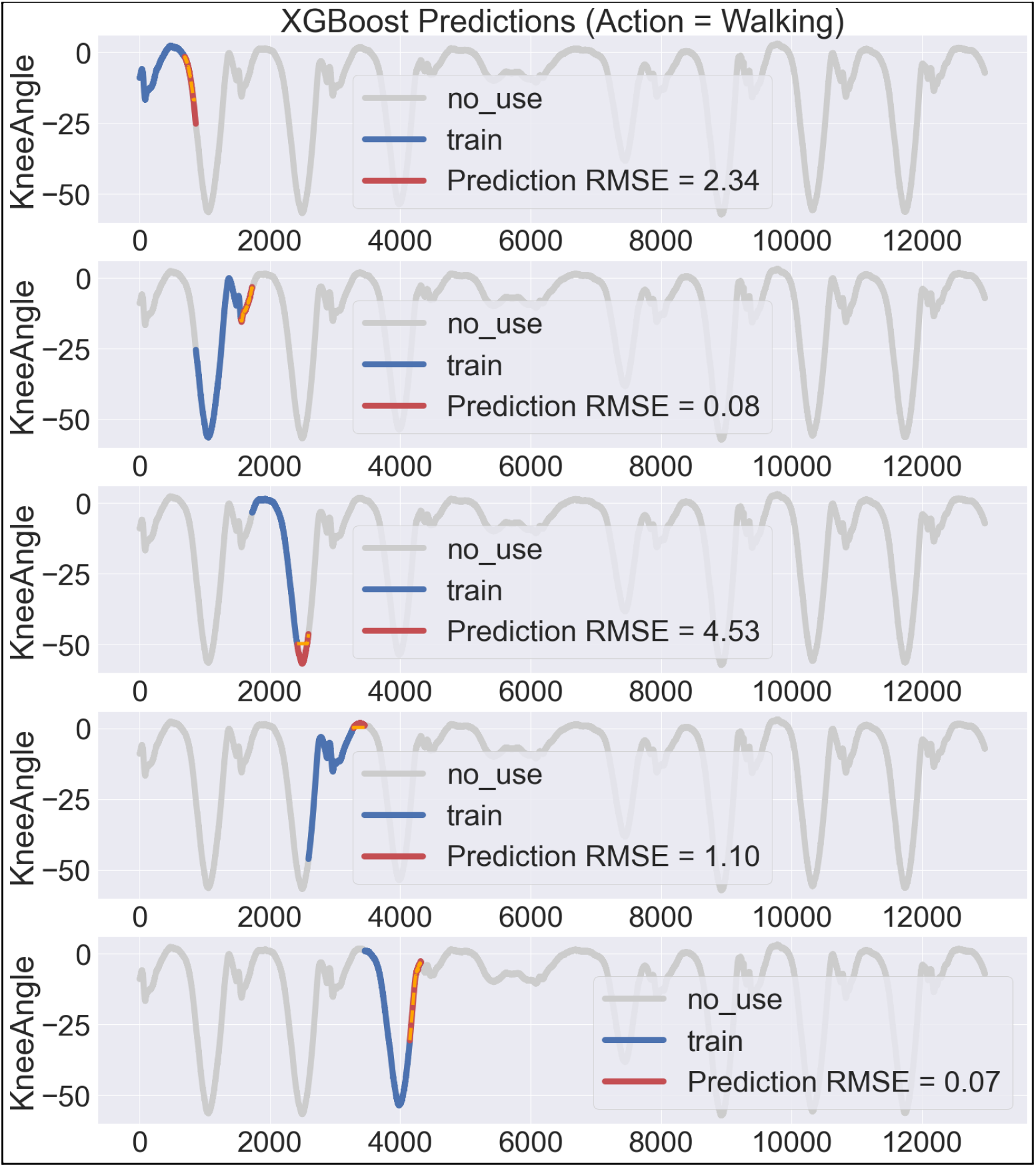
80% training and 20% test predictions using XGBoost for a single subject on a rolling start cross validation

